# Impact of the WHELD/Brief Psychosocial Therapy intervention on psychosis in people with dementia: A Cluster Randomized Trial

**DOI:** 10.1101/2024.09.12.24313538

**Authors:** C Ballard, J McDermid, K Mills, A Sweetnam, J Fossey

## Abstract

Neuropsychiatric Symptoms (NPS), particularly psychosis, are common in dementia and can significantly impact patient outcomes, caregivers and disease trajectory. Psychosis, which includes hallucinations and delusions, occurs in up to 50% of people with dementia and has been linked with lower quality of life and faster cognitive decline. While best practice guidelines have highlighted the importance of non-pharmacological treatments for NPS, evidence-based non-pharmacological approaches are limited. This exploratory analysis of a cluster randomized control trial (RCT) from the WHELD programme compares the WHELD/Brief Psychosocial Therapy intervention with treatment as usual in a 9-month trial across 69 UK nursing homes (N=8477, 553 completed). The current report analyzed outcomes for the participants with dementia-related psychosis (N=163) participating in the trial. Whilst the WHELD/Brief Psychosocial Therapy intervention did not significantly reduce NPI psychosis score, it did significantly improve apathy (p=0.006), agitation (p=0.038) and quality of life (p=0.01) in participants with psychosis. In addition there was a non-significant numerical improvement in caregiver perceived disruptiveness. These findings suggest that whilst the WHELD/ Brief Psychosocial Therapy intervention does not directly alleviate psychosis in people with dementia, it does significantly improve related neuropsychiatric symptoms and quality of life, offering meaningful benefits to people with dementia experiencing distressing psychotic symptoms.

## Introduction

Neuropsychiatric symptoms (NPS) occur in almost all people with dementia at some point during their illness (Aalten et al 2007). These symptoms are distressing for people with dementia and their family and professional caregivers and are associated with a detrimental impact on disease course and outcomes (Ismail et al 2022). Psychosis is a key neuropsychiatric domain, consisting of hallucinations, delusions, and delusional misidentification; with an estimated prevalence of 50% over the disease course and which impacts specifically on accelerated decline, quality of life, and institutionalization (Ismail et al 2022). Concurrent NPS such as agitation, depression, and apathy commonly co-occur with psychosis and contribute significantly to the detrimental impact on quality of life (Choi et al 2022)…

Best practice guidelines advocate non-pharmacological treatments for NPS before pharmacological treatments are considered (eg CSM 2012). This is however problematic for the treatment of psychosis, as there are very limited options for evidence based non-pharmacological approaches.

In the current paper, we report an additional analysis of a factorial randomized control trial (RCT) completed as part of the WHELD nursing home programme. WHELD combines person centred care, anti-antipsychotic review and enjoyable activities with social interaction (utilizing the Brief Psychosocial Therapy programme). Reported benefits in treating agitation include a reduction in the use of anti-psychotics and other psychotropic medication, along with improvements in the quality of life. These outcomes were observed across 4 main RCTs comparing WHELD (in person or through a digitalized version of the programme with virtual coaching) to standard treatment practices. In this report, we specifically evaluate the impact of the Brief Psychosocial Therapy Module of WHELD on psychosis, concurrent NPS and quality of life in people with dementia experiencing psychosis.

## Methods

We completed a 9-month cluster-randomized trial in 69 nursing homes in the UK. The study received full ethical approval from the South-Central Oxford Research Ethics Committee. The trial is registered (ISRCTN62237498) and a fuller description of the methods and the primary results have been published (Ballard et al, 2018). The current paper presents an exploratory analysis focussing on participants within the study who were experiencing dementia-related psychosis.

### Participants

The participants all met criteria for dementia (Clinical Dementia Rating (Morris, 1993) (score of 1 or greater) and the Functional Assessment Staging (Reisberg, 1988) (stage 4 or greater). All residents meeting these broad inclusion criteria were invited to take part. Procedures for consent and assessment of mental capacity are described fully elsewhere (Ballard et al, 2016).

### Interventions

The nursing homes allocated to the WHELD intervention received a person centred care training and implementation programme, which also incorporated antipsychotic review and Brief Psychosocial Therapy to augment enjoyable activities and social interaction with and without exercise The interventions were delivered by trained WHELD therapists, supported by a minimum of 2 WHELD champions in each participating nursing home.

#### Person-centered care

The person-centered care intervention was based on the Focussed Intervention for Training of Staff (FITS) program, which demonstrated significant benefit in a robust randomized controlled trial (Fossey et al, 2016).

#### Antipsychotic review

Antipsychotic review focused on antipsychotic prescriptions by primary care physicians or psychiatry specialists, based on guidelines from the National Institute for Health and Clinical Excellence dementia guidelines (National Institute for Health and Clinical Excellence, 2006) and antipsychotic guidance developed by the Alzheimer’s Society in partnership with the Department of Health in England. The intervention combined training within the nursing homes to understand the issues related to antipsychotic use in these individuals, and to trigger antipsychotic reviews of residents already receiving antipsychotics. This was supported with parallel primary care workshops.

#### Brief Psychosocial Therapy (promoting social interaction with enjoyable activities)

A manual for the social interaction intervention was developed to operationalize the way social activities were selected, to enhance resident interactions with staff, family, and volunteers, and increase the amount of time residents spent in enjoyable activities, based on the Brief Psychosocial Therapy Programme. The objectives were to ensure planned enjoyable activities with social interactions for each resident at least three times per week, aiming for residents to have a minimum of 60 minutes a week of enjoyable activity with social interaction.

#### Exercise

The exercise intervention aimed to increase exercise through enjoyable physical activities based on the Seattle protocols (Teri et al, 2008) and NEST manual (Buettner & Fitzsimmons, 2009), aiming to achieve at least 1 hour a week of enjoyable exercise.

### Outcome Measures

Psychosis, depression and apathy were assessed using the relevant domains of the Neuropsychiatric Inventory, nursing home version (Cummings et al, 1994) (domains for visual hallucinations and delusions – combined as a psychosis score and apathy). Agitation was evaluated by informant interview using the 29-item Cohen-Mansfield Agitation Inventory (Cohen-Mansfield et al, 1989). Quality of life (QoL) was measured by the DEMQOL-Proxy (Mulhern et al 2013), a 31-item informant questionnaire.

Research assistants completed the assessments at baseline and at a 9 month follow-up, blind to intervention allocation.

### Randomization

Randomization was undertaken at the North Wales Organisation for Randomised Trials in Health, using R (statistical package) (Russel et al, 2011). Nursing homes were contacted in the order of appearance on a randomized list. All recruitment and baseline assessments were completed prior to randomization.

### Sample Size

The primary outcome in the original study was quality of life, measured with the DEMQol Proxy. The original trial was powered for a 0.3 effect size, assuming an intra-home correlation of 0.05, to achieve 90% power to detect a difference between groups to the 0.05% level of significance; requiring a sample size of approximately 840 participants to allow for drop-outs… The current analysis is exploratory, and there was hence to specific power calculation undertaken.

### Statistical Analysis

The main outcome measure for the current exploratory analysis was NPI psychosis (combining the total of the NPI hallucinations and delusions domains) in participants scoring >1 on a combined score for NPI hallucinations and NPI delusions. Further exploratory analyses were conducted focussing on agitation, apathy, depression, impact on caregivers and quality of life in participants with psychosis at baseline. All analyses used the multilevel modelling approach to ANOVA, with the value at 9 months as the response and the baseline value as the covariate. The treatment group was the participants receiving the WHELD/Brief Psychosocial Therapy Intervention, the comparison group received treatment as usual..

## Results

### Cohort Characteristics

Sixty nine nursing homes with 847 participants were randomized, of whom 553 (65%) completed the study. The mean age of participants was 88.5 (SE 0.53), 628 (74%) were female and 686 (81%) had had moderately severe or severe dementia based on the FAST score.

### Outcomes

163 (59%) of the participants completing the study scored one or more on the combined NPI psychosis score. In this group, there were no significant benefits in NPI psychosis score between the group receiving and not receiving the WHELD/Brief Psychosocial Therapy intervention. However, WHELD/ Brief Psychosocial Intervention did confer significant benefits in the severity of apathy (NPI domain G) and agitation (29 item CMAI); and there was a non-significant numerical advantage for the group receiving the Brief Psychosocial Therapy intervention on the NPI total disruptiveness score. The results are shown in more detail in table 1.

|  | F | P |
| --- | --- | --- |
| NPI psychosis score | 0.25 | 0.62 |
| NPI Apathy score | 7.7 | 0.006*** |
| CMAI score | 4.4 | 0.038* |
| NPI -Total Disruptiveness score | 3.7 | 0.057 |
| DEMqol proxy Quality of Life | 6.15 | 0.01** |

\*P<0.05, \*\* p≤0.01, \*\*\* p≤0.001

### WHELD Factorial RCT

In a further confirmatory analysis, we also examined the 79 patients with dementia related psychosis participating in an earlier factorial RCT of 277 people with dementia in 16 UK nursing homes. The Personalized activity/Brief Psychosocial Therapy intervention did not confer any direct benefits to psychotic symptoms, but there was a significant benefit in concurrent agitation (F=10.3, P=0.003). In addition, there was a significant benefit in the Personalized activity/Brief Psychosocial Therapy group in quality of life, as measured by the DEMQOL proxy (F= 6.9 P=0.01).

## Discussion

Psychosis is a common and distressing symptom amongst people with dementia, particularly amongst those living in care homes. The current evaluation confirms a frequency of either delusions and/or hallucinations of almost 30%. In the current evaluation from the WHELD cluster RCT, the WHELD/Brief Psychosocial Therapy intervention conferred significant benefits in people with psychosis compared to treatment as usual. Although there was no direct benefit in presenting psychotic symptoms, key concurrent neuropsychiatric symptoms such as apathy and agitation did significantly improve, there was a significant benefit in quality of life amongst participants with psychosis and there was an associated numerical benefit in the disruptiveness experienced by the professional caregivers. Additionally, further analysis demonstrated similar benefits in agitation and quality of life for people with dementia related psychosis from the previous WHELD factorial study (Ballard et al 2016)..

This is a potentially important step forward, as to date there is very limited evidence that any non-pharmacological interventions benefit people with dementia-related psychosis. Recent studies have also highlighted the frequency and impact of concurrent neuropsychiatric symptoms on the quality of life in people with dementia (Choi et al, 2022), highlighting the broader benefits of improving these symptoms, which was reflected in the benefits in quality of life that were evident in the participants with dementia related psychosis in the current study. These results highlight that non-pharmacological interventions focussing on personalized activities can offer significant benefits for people with dementia experiencing psychosis, although further work is also imperative to develop more targeted interventions that also confer direct benefits for the psychotic symptoms.

The WHELD study was a robust cluster RCT, but it should be acknowledged that this was a post-hoc exploratory analysis, and needs to be interpreted cautiously. Nevertheless these results are encouraging. The Brief Psychosocial Therapy module within WHELD, promoting personalized activities, has the advantage of being quick and straightforward to deliver for individual patients. It follows an established and well-operationalized protocol and has the potential for widespread implementation.

## Data Availability

All data produced in the present study are available upon reasonable request to the authors

